# PhenomAD-NDD: the Phenomics Aggregation Database of comorbidities in 51,227 pediatric individuals with NeuroDevelopmental Disorders

**DOI:** 10.1101/2023.11.29.23299167

**Authors:** Alexander J M Dingemans, Sandra Jansen, Jeroen van Reeuwijk, Nicole de Leeuw, Rolph Pfundt, Janneke Schuurs-Hoeijmakers, Bregje W van Bon, Carlo Marcelis, Charlotte W Ockeloen, Marjolein Willemsen, Pleuntje J van der Sluijs, Gijs W E Santen, R Frank Kooy, Anneke T Vulto-van Silfhout, Tjitske Kleefstra, David A Koolen, Lisenka E L M Vissers, Bert B A de Vries

## Abstract

The prevalence of comorbidities in individuals with neurodevelopmental disorders (NDD) is not well understood, while these are important for accurate diagnosis and prognosis in routine care and for characterizing the clinical spectrum of NDD syndromes. Therefore, we developed PhenomAD-NDD: an aggregated database with comorbid phenotypic data of 51,227 individuals with NDD, all harmonized into Human Phenotype Ontology (HPO), with in total 3,054 unique HPO terms. We demonstrate that almost all congenital anomalies are more prevalent in the NDD population than in the general population and the NDD baseline prevalence allows for approximation of enrichment of symptoms. Such analyses for 33 genetic NDDs for instance shows that 32% of enriched phenotypes is currently not reported in the clinical synopsis in OMIM. PhenomAD-NDD is open to all via a visualization online tool and allows to determine enrichment of symptoms in NDD.

## 1 Introduction

In 1938, Penrose was the first to describe the clinical features of individuals with neurodevelopmental disorders (NDD) in *A Clinical and Genetic Study of 1280 Cases of Mental Defect* [1]. Although he mainly focused on inheritance patterns and looking at nature versus nurture effects, he also reported congenital malformations in this cohort. Despite almost a century of additional research, of which many reported on associated clinical features in individuals with NDD [2–15], only the prevalence of comorbid mental health disorders and epilepsy (respectively up to 40% and 26%) have been determined [16, 17]. For most specific phenotypic characteristics the overall prevalence, however, eludes us, although these additional phenotypic features are essential when taking care of a child with developmental delay (DD) or intellectual disability (ID). These comorbid clinical features assist in making a clinical diagnosis, are crucial for adequate variant interpretation and provide information on prognosis and treatment. NDD affect 2-3% of the population [18], and the absence of information on comorbidities can thus be perceived as more than just a general clinical problem. Especially following the introduction of next-generation sequencing (NGS), which allowed to find more genetic diagnoses for individuals with rare disease, and NDD in particular, there is a progressive need for more accurate and quantitative phenotypic data for NDD. With the rise of NGS in clinic, the ‘Genome Aggregation Database (gnomAD) Consortium’ made headway at unprecedented scale by providing an essential contribution to the field which captures the extent of genetic variation in >120k individuals, which today is used as a rich resource for assessing and prioritizing genetic variation in health and disease [19]. In part, the successes of gnomAD relied on standardization and aggregating data set of thousands of individuals [20]. Whereas advances have been made to allow for clinical phenotypes to be captured in standardized terminology, such as human phenotype ontology (HPO) [21, 22], phenomics data is mostly only captured at the individual patient level, with some exceptions such as DECIPHER [23]. This prohibits not only systematic large-scale analyses, such as for instance on the prevalence and existence of comorbidities in a given patient population, but also hampers accurate genotype-phenotype correlations as without such a cohort prevalence, one cannot be confident that a particular anomaly is part of a specific disorder. In this study, we therefore set out to describe the baseline prevalence of comorbid features in pediatric individuals with NDD by aggregating our inhouse clinical data with a systematic review of all available literature related to the pediatric clinical spectrum of NDD. These data and its infrastructure are freely available in PhenomAD-NDD, an online repository, which will not only be essential to determine what features are enriched for a particular genetic disorder, but will also improve our general understanding of NDD and associated comorbidities.

## 2 Results

### 2.1 Overview of PhenomAD-NDD

We included 1,477 individuals with NDD seen at the Radboud university medical center in PhenomAD-NDD, of whom 969 (66%) were male, and 508 (34%) were female, and with a median age of 8.8 years (IQR 7.2 years). A literature review strategy yielded 9,919 records reporting on NDD and associated comorbidities, which after exclusion of duplicates (n=3,605) left 5,314 for manual screening (Supplementary Table 1). Two hundred ninety-one were deemed possible inclusions, which after assessment of eligibility, resulted in 38 included studies. Subsequent screening of the references identified another 27 studies for inclusion, bringing the total to 65 studies (Figure 1). Extracting the phenotypic data and converting these to HPO terms provided data for 49,750 children with NDD. Collectively, the total number of individuals included in PhenomAD-NDD was 51,227, with 3,054 unique HPO terms, and on average 2.5 HPO terms per patient (12.5 when including parent-HPO-nodes). The HPO terms per individual for the individuals seen in Nijmegen were relatively normally distributed, indicating no apparent clustering of congenital anomalies in specific individuals (Supplemental Figure 2).

**Figure 1:**
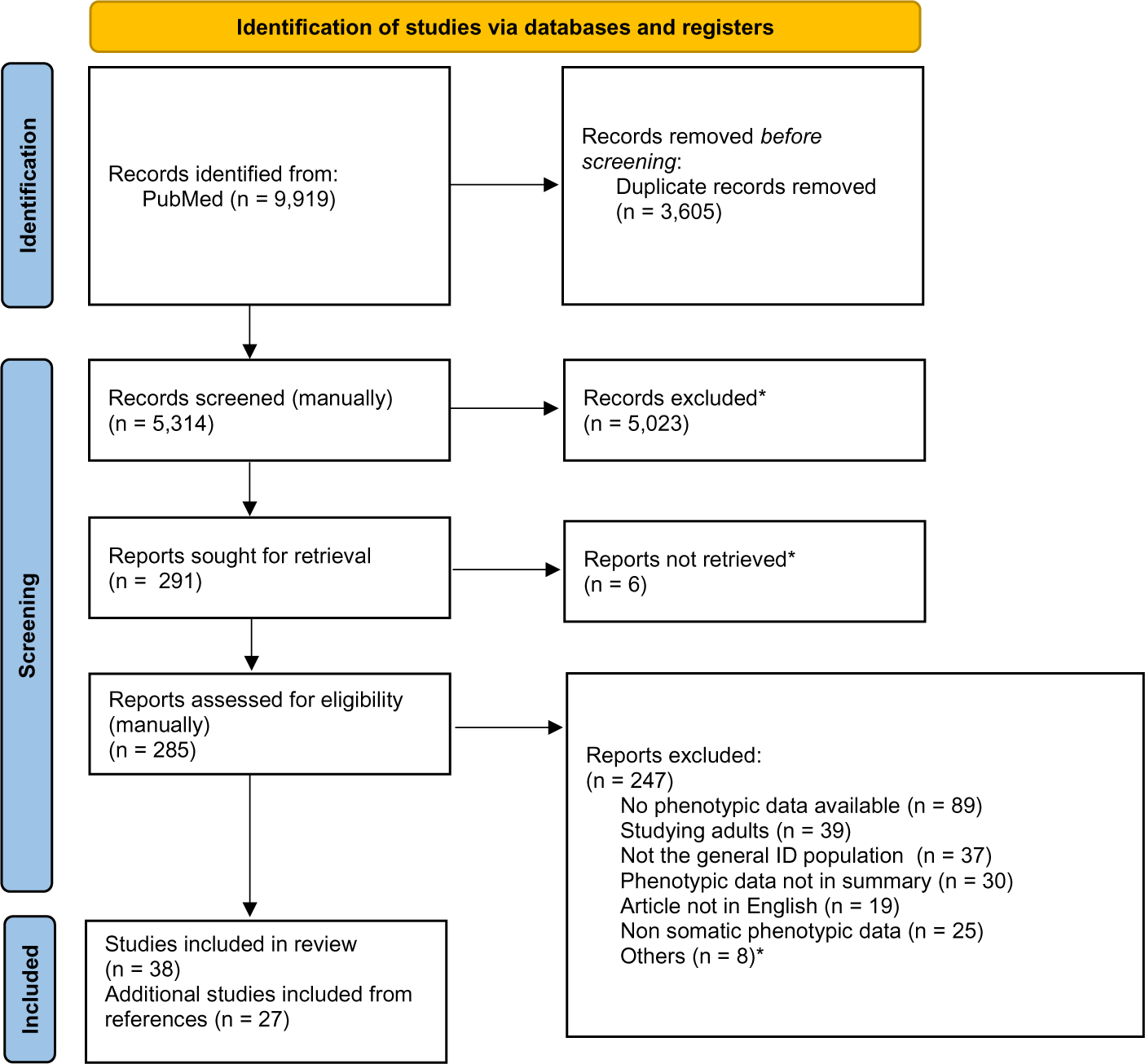
PRISMA 2020 flow diagram which shows the complete search strategy utilized in this study [38]. *For the reason for exclusion per study, please see supplementary table 1.

**Figure 2:**
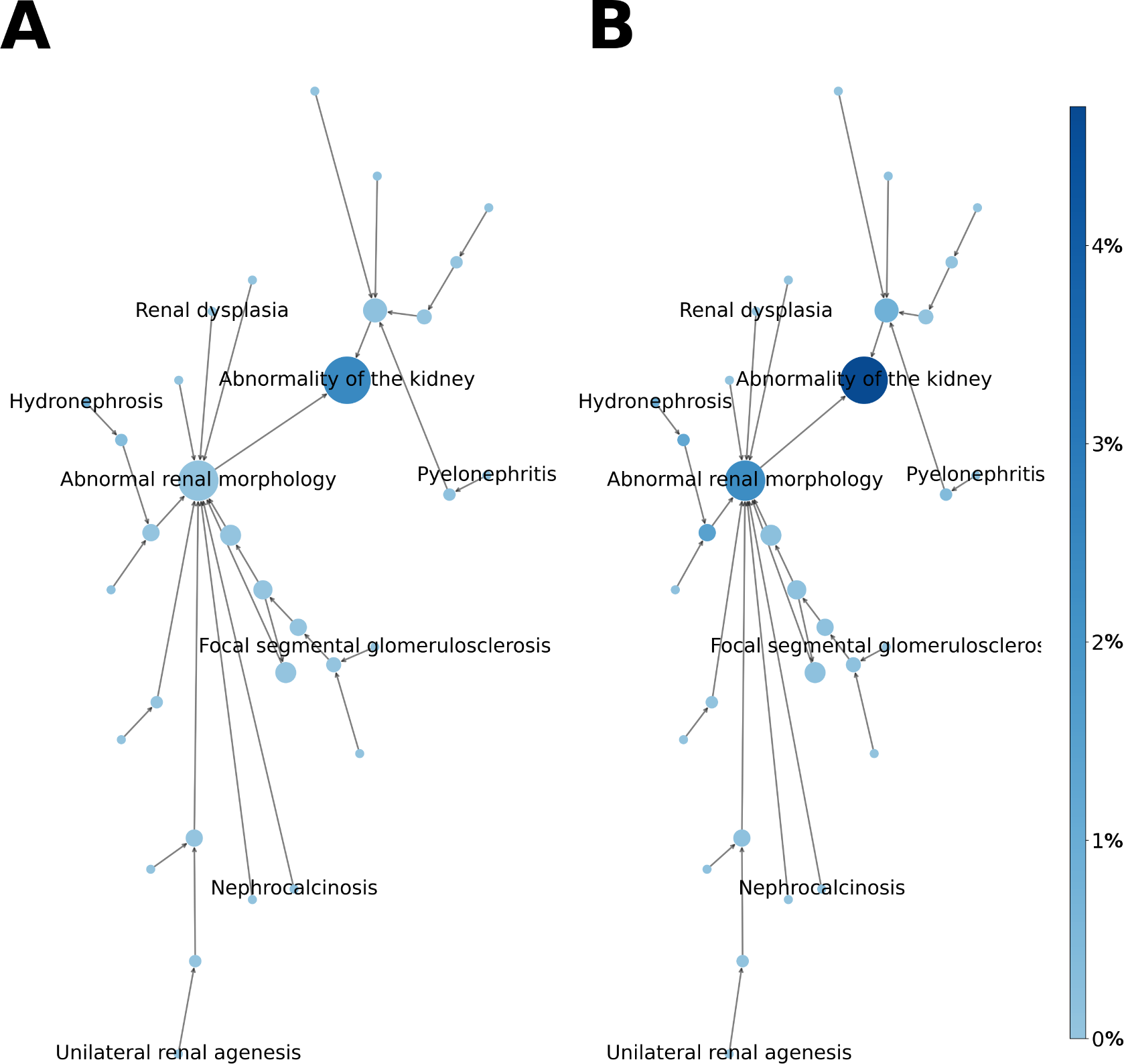
The prevalence of kidney-related HPO terms in PhenomAD-NDD (in %). A) shows the prevalence of only that specific symptom; although B) shows the same HPO terms, now the HPO terms are counted as positive if one of the child nodes is positive - to minimize interobserver variability. The difference when looking at, for instance, “Abnormal renal morphology” is quite significant (0.1% vs. 2.3%).

Looking at the contents of PhenomAD-NDD, epilepsy (21.26% [20.55% - 22.00%]) and behavioral problems (44.38% [43.47% - 45.30%]) were relatively common (Table 1). Autistic behavior was noted in a fifth (21.34% [20.39% - 22.33%]) of individuals, while delayed speech (52.02% [49.91% - 54.19%]) was as prevalent as a delay in the development of motor skills (51.93% [50.35% - 53.54%]). Some individuals had some abnormality of the musculoskeletal system (25.77% [25.19% - 26.36%]) mainly scoliosis (28.54% [26.75% - 30.42%]) or joint hypermobility (13.5% [11.73% - 15.55%]). Morphological anomalies of the kidney (2.3% [1.59% - 3.22%]) and the heart (7.6% [7.09% - 8.13%]) were relatively few. All individuals had a neurodevelopmental abnormality, with the vast majority (91.89% [90.83% - 92.96%]) diagnosed with intellectual disability. For 25.8% of these individuals, the severity of the ID was known, and over a third (44.81% [43.30% - 46.37%]) of this subgroup was diagnosed with mild ID, while 24.99% [23.87% - 26.16%] had moderate ID, 19.62% [18.62% - 20.65%] had severe ID diagnosed and 10.57% [9.85% - 11.34%] had profound ID. Less than a quarter of our study population reported immunological issues (12.6% [11.95% - 13.26%]), which were more common than endocrinological problems (3.35% [2.77% - 4.01%]). The complete overview of all clinical features in PhenomAD-NDD can be found online and in the electronic supplement of this paper (Figure 3).

**Figure 3:**
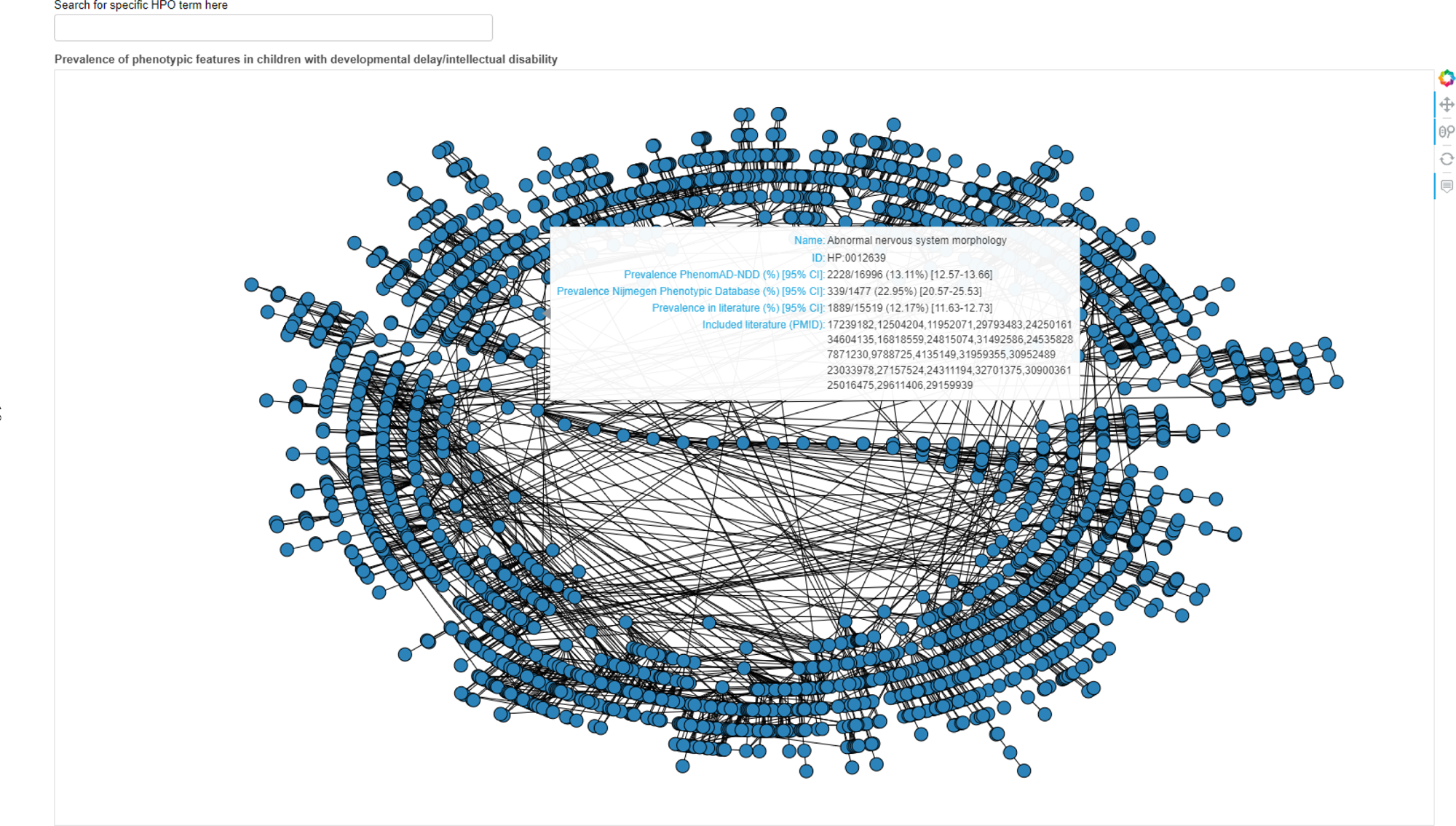
A screenshot of one of the two produced tools: this one is developed with the Bokeh [39] library, which can be used to view the prevalence of any clinical feature in PhenomAD-NDD. A specific symptom can be looked up, which produces the prevalence of that specific HPO term including any child nodes as well. The tool is available as a supplement to this paper and on the Human Disease Genes website series(https://humandiseasegenes.nl/phenomadndd/)[37] and can be used as the definition of the baseline somatic comorbidity in pediatric individuals with NDD.

**Table 1:**
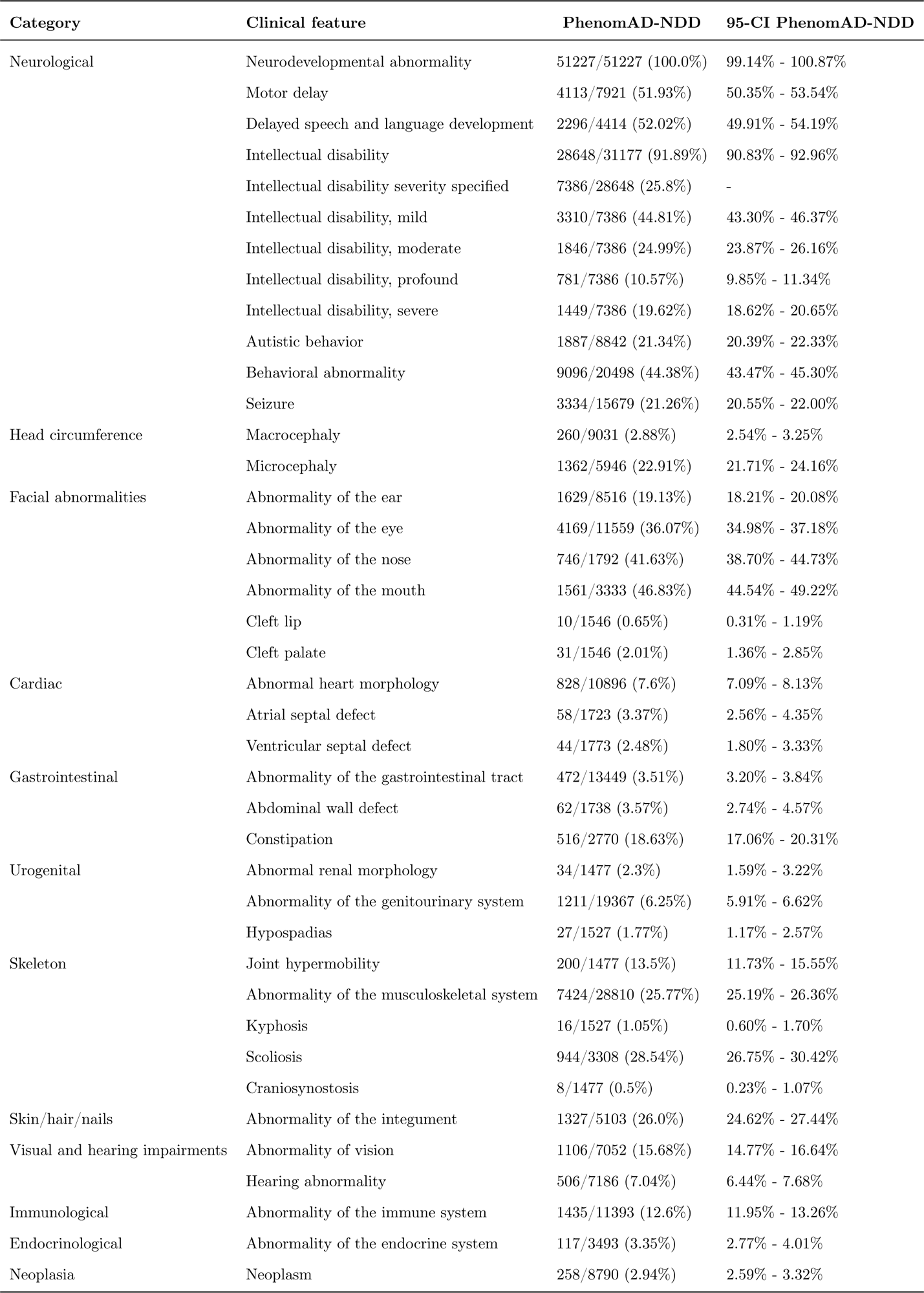
The prevalence of some standard clinical features in the 51,227 individuals included in PhenomAD-NDD.

### 2.2 Subgroup analysis

When investigating several subgroups in our novel clinical cohort of 1,477 individuals for differences between sexes for instance, behavioral abnormalities (69% vs 55%, *p* <0.00001), autistic behavior (39% vs 21%, *p* <0.00001) and urogenital problems (22% vs 10%, *p* <0.00001) were all more common in boys than in girls, respectively. However, most clinical features, such as ID and its severity, were not statistically significantly different between the sexes (Figure 4). When looking at the phenotype of individuals with different severity of ID, seizures were significantly more common in the severe/profound ID group when compared to the mild/moderate ID group (48% vs 14%, *p* <0.00001). In comparison, behavioral abnormalities were more present in the mild/moderate ID group than in the severe/profound ID group (78% vs. 58%, *p*=0.0004).

**Figure 4:**
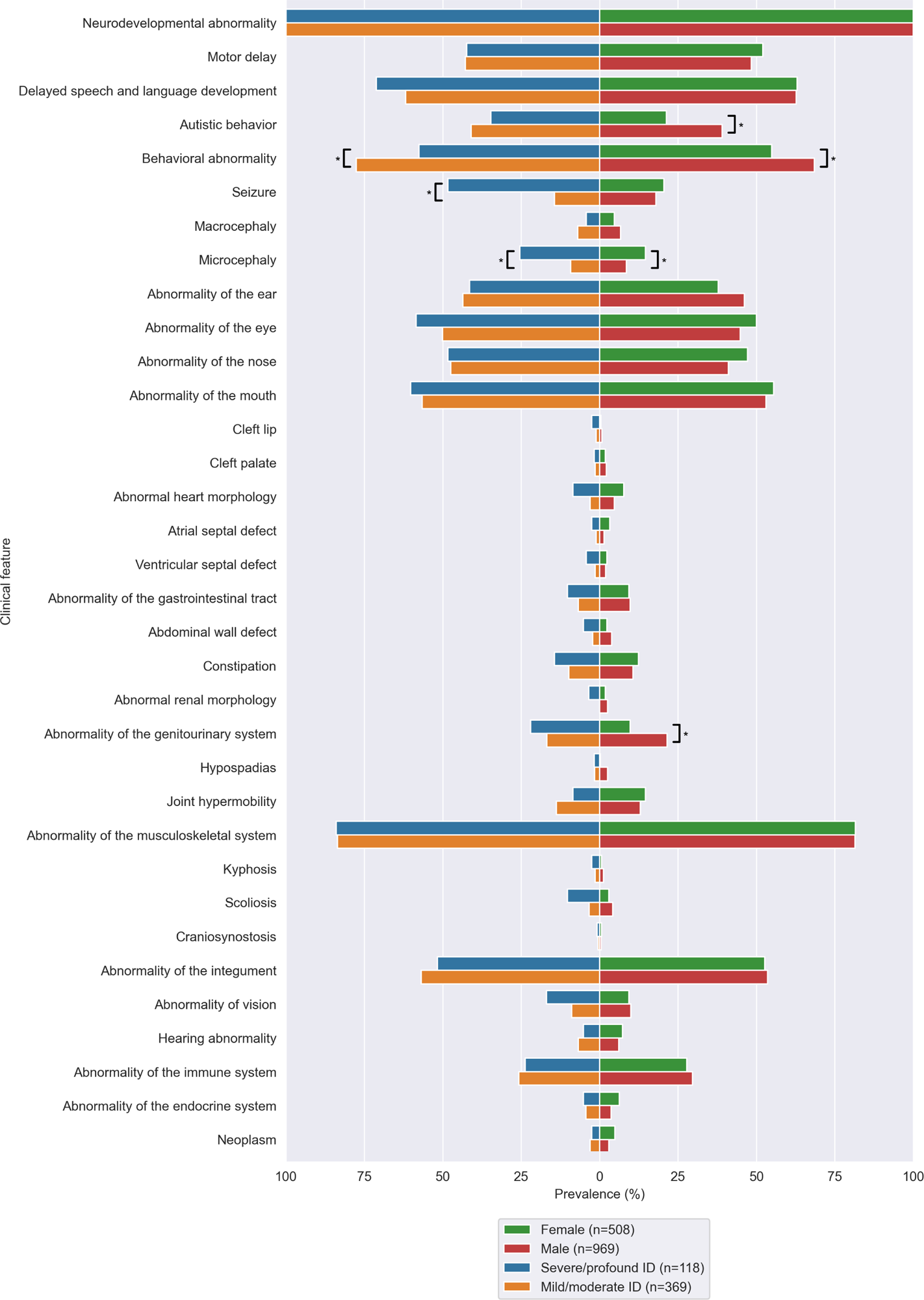
Two subgroups are compared within the novel cohort, based on sex and severity of intellectual disability - when reported. Autistic behaviour and moderate intellecutal disability are more common in males, while congenital heart defects are more prevalent in females. Next to that, individuals with more severe ID report more microcephaly, seizures and congenital heart defects. * = statistically significant at the Bonferroni corrected level

### 2.3 Comparison to EUROCAT

To determine whether congenital anomalies were more present in the NDD population when compared to the general population, the European network of population-based registries for the epidemiological surveillance of congenital anomalies (EUROCAT) was consulted. It collects prevalence information on congenital anomalies in the general population [24–26] and was therefore suitable as a control group. In total, data of 11,616,332 individuals of EUROCAT, collected from 37 institutions of 18 countries were included in these analyses (Supplemental Figure 1). These provide an extensive overview of the baseline prevalence of congenital anomalies in all births: congenital heart defects for instance are present in 0.7%, with ventricular septal defects accounting for roughly half— while the prevalence of these in individuals with NDD is 10 times higher (7.6%). Hypospadias are present in 0.2% of individuals in EUROCAT, and individuals with NDD report this symptom ten times more often as well (1.8%). For abdominal wall defects, the difference between EUROCAT and NDD is almost 100-fold (0.04% vs 3.6%, respectively).

All congenital anomalies were present in less than 1% of individuals in EUROCAT and almost all investigated congenital anomalies were present more frequently in our NDD cohort when comparing the prevalence to EUROCAT (Table 2). Some abnormalities, like congenital glaucoma and Hirschsprung’s disease, were not reported at all in PhenomAD-NDD: for the 40 congenital anomalies that were reported in both PhenomAD-NDD and EUROCAT, all were more prevalent in PhenomAD-NDD, with 34 reaching statistical significance (85%) after Bonferroni correction. Furthermore, while PhenomAD-NDD exhibited a higher frequency of almost all congenital anomalies when compared to EUROCAT, it is important to acknowledge the potential ascertainment bias inherent to our findings. Certain anomalies may be less likely to come to clinical attention unless a more thorough clinical evaluation is performed, often triggered by the neurodevelopmental condition.

**Table 2:** The prevalence of congenital anomalies in our NDD cohort compared with the prevalence of these anomalies in the EUROCAT registry.

| Anomaly | HPO ID | Prevalence PhenomAD-NDD (%) | Prevalence Eurocat (%) | p-value |
| --- | --- | --- | --- | --- |
| Abdominal wall defect | HP:0010866 | 3.567 | 0.037 | 0.000000e+00 |
| Anencephalus and similar | HP:0002323 | 0.000 | 0.003 | 1 |
| Annular pancreas | HP:0001734 | 0.000 | 0.002 | 1 |
| Anophthalmos | HP:0000528 | 0.062 | 0.001 | 1.467373e-03 |
| Anophthalmos/microphthalmos | HP:0000568; HP:0000528 | 0.406 | 0.008 | 1.727896e-138 |
| Ano-rectal atresia and stenosis | HP:0002023; HP:0002025 | 0.034 | 0.026 | 1 |
| Anotia | HP:0009892 | 0.000 | 0.008 | 1 |
| Aortic atresia/interrupted aortic arch | HP:0012304; HP:0011611 | 0.034 | 0.004 | 2.270432e-01 |
| Aortic valve atresia/stenosis | HP:0010883; HP:0001650 | 0.169 | 0.013 | 1.332385e-11 |
| Arhinencephaly/holoprosencephaly | HP:0001360 | 0.068 | 0.003 | 2.773007e-02 |
| Atresia of bile ducts | HP:0005912 | 0.000 | 0.003 | 1 |
| Atresia or stenosis of other parts of small int... | HP:0012848; HP:0012739 | 0.000 | 0.008 | 1 |
| Atrial septal defect | HP:0001631 | 3.366 | 0.186 | 7.768723e-202 |
| Atrioventricular septal defect | HP:0006695 | 0.406 | 0.032 | 2.353218e-13 |
| Bilateral renal agenesis including Potter syndrome | HP:0010958 | 0.000 | 0.003 | 1 |
| Bladder exstrophy and/or epispadia | HP:0002836 | 0.000 | 0.005 | 1 |
| Choanal atresia | HP:0000453 | 0.000 | 0.008 | 1 |
| Cleft lip | HP:0000202 | 1.906 | 0.079 | 3.013225e-203 |
| Cleft palate | HP:0000175 | 2.005 | 0.056 | 7.862696e-220 |
| Club foot - talipes equinovarus | HP:0001762 | 1.422 | 0.056 | 1.267641e-102 |
| Coarctation of aorta | HP:0001680 | 0.339 | 0.036 | 5.895378e-08 |
| Common arterial truncus | HP:0001660 | 0.135 | 0.005 | 4.249057e-08 |
| Congenital cataract | HP:0000519 | 0.135 | 0.013 | 2.934049e-03 |
| Congenital constriction bands/amniotic band | HP:0009775 | 0.000 | 0.002 | 1 |
| Congenital glaucoma | HP:0008007; HP:0008041 | 0.000 | 0.003 | 1 |
| Congenital Heart Defects | HP:0001627 | 7.599 | 0.710 | 0.000000e+00 |
| Congenital hydronephrosis | HP:0000126 | 0.948 | 0.140 | 1.877582e-15 |
| Craniosynostosis | HP:0001363 | 0.542 | 0.024 | 1.199291e-33 |
| Cystic adenomatous malf of lung § | HP:0010959 | 0.000 | 0.008 | 1 |
| Diaphragmatic hernia | HP:0000776 | 0.135 | 0.021 | 3.452795e-02 |
| Double outlet right ventricle § | HP:0001719 | 0.000 | 0.010 | 1 |
| Duodenal atresia or stenosis | HP:0002247; HP:0100867 | 0.000 | 0.012 | 1 |
| Ebstein's anomaly | HP:0010316 | 0.000 | 0.004 | 1 |
| Encephalocele | HP:0002084 | 0.000 | 0.004 | 1 |
| Gastroschisis | HP:0001543 | 0.000 | 0.023 | 1 |
| Hip dislocation and/or dysplasia | HP:0002827; HP:0001385 | 1.557 | 0.063 | 5.096832e-225 |
| Hirschsprung's disease | HP:0002251 | 0.064 | 0.014 | 5.469745e-01 |
| Hydrocephalus | HP:0000238 | 2.145 | 0.031 | 0.000000e+00 |
| Hypoplastic left heart | HP:0004383 | 0.068 | 0.015 | 5.386133e-01 |
| Hypoplastic right heart | HP:0010954 | 0.000 | 0.003 | 1 |
| Hypospadias | HP:0000047 | 1.768 | 0.175 | 3.237488e-48 |
| Indeterminate sex | HP:0000062 | 0.068 | 0.005 | 1.063194e-01 |
| Limb reduction defects | HP:0009815 | 12.796 | 0.038 | 0.000000e+00 |
| Microcephaly | HP:0000252 | 22.906 | 0.024 | 0.000000e+00 |
| Mitral valve anomalies | HP:0001633 | 0.135 | 0.004 | 4.930881e-10 |
| Multicystic renal dysplasia | HP:0000003 | 0.000 | 0.029 | 1 |
| Neural Tube Defects | HP:0410043 | 0.475 | 0.026 | 1.654476e-130 |
| Oesophageal atresia with or without tracheo-oes... | HP:0002032 | 0.068 | 0.023 | 7.754114e-01 |
| Omphalocele | HP:0001539 | 0.203 | 0.013 | 7.472370e-08 |
| Oro-facial clefts | HP:0002006 | 7.893 | 0.135 | 0.000000e+00 |
| PDA as only CHD in term infants (>=37 weeks) | HP:0001643 | 1.151 | 0.035 | 3.944004e-108 |
| Polydactyly | HP:0010442 | 0.406 | 0.087 | 2.121606e-04 |
| Posterior urethral valve and/or prune belly | HP:0010957; HP:0004392 | 0.000 | 0.009 | 1 |
| Pulmonary valve atresia | HP:0010882 | 0.000 | 0.009 | 1 |
| Pulmonary valve stenosis | HP:0001642 | 0.609 | 0.042 | 8.074645e-24 |
| Single ventricle | HP:0001750 | 0.000 | 0.004 | 1 |
| Situs inversus | HP:0011616; HP:0011538;<br>HP:0003363; HP:0001696;<br>HP:0031592; | 0.000 | 0.005 | 1 |
| Skeletal dysplasias § | HP:0002652 | 0.000 | 0.005 | 1 |
| Spina Bifida | HP:0002414 | 0.475 | 0.019 | 7.040170e-174 |
| Syndactyly | HP:0001159 | 6.892 | 0.044 | 0.000000e+00 |
| Tetralogy of Fallot | HP:0001636 | 0.393 | 0.029 | 3.021266e-14 |
| Total anomalous pulm venous return | HP:0005160 | 0.000 | 0.006 | 1 |
| Transposition of great vessels | HP:0001669 | 0.339 | 0.031 | 1.934738e-09 |
| Tricuspid atresia and stenosis | HP:0011662; HP:0010446 | 0.000 | 0.005 | 1 |
| Ventricular septal defect | HP:0001629 | 2.482 | 0.338 | 5.225090e-53 |

### 2.4 Enrichment analysis

To detect phenotypic features that are predictive of a particular genetic syndrome and demonstrate the additional power of PhenomAD-NDD, the enrichment of clinical features was calculated and compared to the HPO terms in the clinical synopsis of OMIM for 33 NDD-related syndromes [27] for which also high-quality data is available in the HDG websites for individuals with a clinical and molecularly (American College of Medical Genetics and Genomics class 4 or 5 variant) proven diagnosis for these 33 disorders (Supplementary table 2). When looking at all enriched symptoms in these 33-NDD-related syndromes with a minimal prevalence of 1%, 1,541 features were enriched according to PhenomAD-NDD: 1,041 (68%) of those were recorded in the clinical synopsis — indicating that 32% of relevant clinical features according to PhenomAD-NDD are currently missing in OMIM. This varies significantly per genetic syndrome, from 16% (MRD50, OMIM #617787) to 100% (NEDHELS, #617171, Figure 5) of concordance of the enriched features with the clinical synopsis. Furthermore, the symptoms that are not present in OMIM were on average enriched by a factor of 8.2 in the genetic NDD related syndromes compared to PhenomAD-NDD.

**Figure 5:**
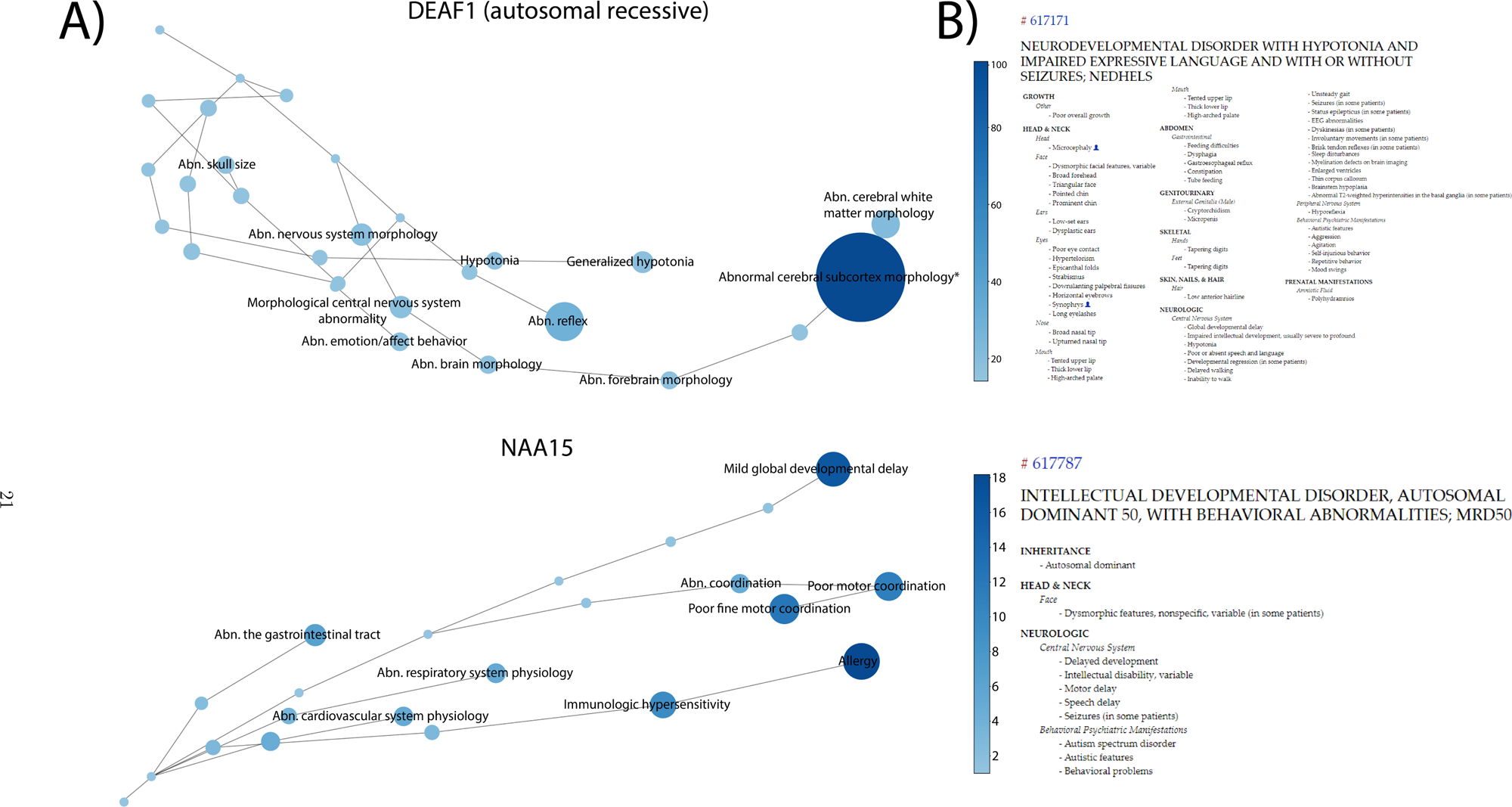
**A)** The results of the top 10 results of the enrichment analysis for two of the 33 investigated genetic syndromes are shown here: MRD50 (OMIM #617787), for which 16% of the symptoms are described in the clinical synopis in OMIM, and NEDHELS (OMIM #617171, 100% concordance). Shown are the 10 clinical features that are the most enriched: i.e. with the highest ratio when dividing the prevalence in the syndrome with the prevalence in PhenomAD-NDD. The size of the nodes in the network corresponds to the prevalence in that specific genetic syndrome. The phenotypic data of the investigated syndromes was gathered from the specific website of the Human Disease Gene website series, with the prevalence rate in the syndrome referring to the proportion of individuals with the specific genetic syndrome who exhibit a particular clinical feature. The color corresponds to the enrichment ratio, which compares the prevalence in the syndrome to the prevalence in PhenomAD-NDD. To avoid spurious enrichments, only clinical features with a prevalence of at least 1% are shown in this figure. Using the developed tool in either the electronic supplement or online at the Human Disease Genes website series(https://humandiseasegenes.nl/phenomadndd/)[37], others are able to determine enriched clinical features and generate these figures for their syndrome of interest. **B)** The clinical synopsis of OMIM [27] as shown for these two genetic syndromes.

We next investigated whether 3,350 clinical features described in both OMIM and the HDG dataset for these 33 NDD-related syndromes are in fact enriched, and thus predictive features for the syndrome. The average enrichment for all these terms is 3.2 — indicating that these clinical features are on average 3 times more present in these syndromes than in the general NDD population. However, 628 (19%) have an enrichment lower than 1.0, indicating that these features might not be enriched in the genetic syndrome. Of these, 59 (2%) reach statistical significance after Bonferroni correction, indicating that these are more prevalent in the general NDD population than in the genetic syndrome. An example of such a clinical feature is feeding difficulties for disorder associated with heterozygous pathogenic variants in *DEAF1* (Vulto-van Silfout-de Vries syndrome, OMIM #615828 [28]). The syndrome is characterized by delayed development, behavioral abnormalities, hypotonia and seizures, with a wide range of symptoms described. OMIM indicates feeding difficulties as part of the phenotype; the prevalence of feeding difficulties in the syndrome is, however, only 27%, against 57% in PhenomAD-NDD (RR 0.47, *p*<0.001).

## 3 Discussion

Although investigations on comorbidities in neurodevelopmental disorders started almost a hundred years ago [1–9], PhenomAD-NDD provides an aggregated detailed phenotypic description of a NDD cohort this size and quality, containing 51,227 individuals with NDD. Several other databases that collect phenotypic data do exists such as the LOVD [29], PhenoDis [30], GPCards [31], DECIPHER [23], and the Monarch Initiative [32]. However, only a limited number of these initiatives focus on NDDs specifically. A nice example of the latter is DECIPHER which over the years has collected data of >30k individuals with developmental disorders [23]. The collection of PhenomAD-NDD is however agnostic to the genetic diagnosis, and provides both the nominator and denominator for all features assessed. This difference is crucial to not only capture the prevalence of features in the broader NDD context, but also to allow to determine the relevance of features when describing the phenotypic spectrum of (novel) syndromes facilitating better insights into prognosis.

In PhenomAD-NDD, there currently is data of on average 2.5 HPO terms per individual (12.0 HPO terms including parent-HPO-nodes). These data, available in detail to all, can be used as a benchmark for future disease-gene discovery studies; by comparing the prevalence of a clinical feature to the prevalence in our cohort, one can determine whether a specific symptom is enriched for that particular patient group. For example, we show that epilepsy is expected in 21.3% of the NDD population, as is autistic behavior in 21.3%: two examples of a clinical feature commonly described as a feature of a particular genetic syndrome. We show that if individuals with a pathogenic variant in a gene report epilepsy or autistic behaviour in roughly 20-25% that these numbers are actually the norm in the general NDD population and therefore should not be noted as a distinctive feature of that disorder. The determination of predictive clinical features is essential for the clinician when considering possible syndromic diagnoses in individuals with NDD. The aggregation of multiple data sources sometimes leads to difficulties: scoliosis for instance, was present in 3.8% of the individuals seen in our outpatient clinic, but in almost 48.5% of individuals in the literature for whom scoliosis was reported – indicating a possible reporting bias in the included papers, and therefore elevating the prevalence in PhenomAD-NDD to 28.5%.

However, the aggregation of all these data provides large benefits as well. In recent years, largescale data sets of human sequence data have become freely accessible online, first with ExAC and more recently with gnomAD [19, 33]. These genomic data assist us in the medical interpretation of genetic variants by illustrating the healthy human genetic variation. Furthermore, these data are used to calculate the intolerance to variants in a specific gene (like the pLI score[34]) which have become essential when discovering new disease genes: a gene, suspected to be associated with a novel genetic syndrome should not have similar (or even identical) diplotypes in the healthy population as in affected individuals [35, 36]. PhenomAD-NDD may as such be considered an equivalent of gnomAD for the interpretation at the phenotypic level of individuals in the broader context of neurodevelopmental disorders. Ultimate synergy may be obtained when both phenotypic and genotypic data at this unprecedented scale become available, with more initiatives developed to collect these data, such as the European ‘1+ Million Genomes’ Initiative. Further incorporating these different data sets and adding additional modalities of patient data to PhenomAD-NDD and investigate further opportunities to unify worldwide existing initiatives on the systematic collection of phenotype-genotype information could further increase the utility of the database. However, since these additional modalities are currently not systematically captured together in a uniform manner in clinical practice, this would require a different manner of capturing these (phenotypic) data.

We utilize the data collected in this study for several analyses: first, proving the data enable the determination of enriched clinical features for 33 genetic NDD-related syndromes. We demonstrate that PhenomAD-NDD is able to recover the phenotypic features that are known in the literature remarkably well. Furthermore, because the enrichment analysis constitutes a quantitative analysis, it is possible to calculate the relative prevalence of the clinical features when comparing those with the general NDD population in PhenomAD-NDD. Comparing these with the current standard in clinical practice, OMIM, shows that the investigated clinical synopses are currently missing 32% of statistically significant enriched clinical features. Furthermore, we demonstrate that these symptoms are on average enriched by a factor of 8.2 — indicating the clinical significance of these missing phenotypic features. Looking at the described symptoms in OMIM, these are generally correct, with only 2% of the clinical features described more prevalent in the genetic syndrome than in the general NDD population according to PhenomAD-NDD, indicating only a small proportion of the clinical features described in OMIM exhibit a higher prevalence in PhenomAD-NDD than in the genetic syndrome population. Examining the concordance of the enriched features according to our enrichment analyses and the clinical synopsis of OMIM, we see that there is a substantial variation between genetic syndromes — as some, such as NEDHELS (OMIM #617171) have much more and detailed phenotypic information than others (such as MRD50, OMIM #617787) in OMIM described. This is probably due to the nature of OMIM: since it relies on manual curation of the medical literature, some pages will be more up-to-date than others.

These analyses demonstrate what the actual predicting clinical features are for these genetic syndromes by quantifying the difference in prevalence. This enables the discovery of predicting clinical features for (novel) genetic syndromes, so that healthcare professionals know which symptoms are characteristic of a specific disorder when seeing an individual with NDD. While we show the results for 33 genetic NDD related disorders, anyone can establish the enriched clinical features for any genetic disorder of interest using the developed tool online or in the electronic supplement. It is important to note here that the data in OMIM is still relevant: it all comes down to the question one is attempting to answer. If you are wondering whether a clinical feature is a part of a genetic syndrome (i.e. the prevalence is elevated when compared to the general population), the clinical synopsis of OMIM provides an excellent overview. However, if you actually want to compare the prevalence of a specific clinical feature to the general NDD population, as for instance is the case in variant interpretation, or describing the characteristic features in a novel syndrome, our data is better suited to provide answers.

Next to these analyses, we confirm our clinical hypothesis that congenital anomalies are more present in pediatric individuals with NDD than in the general population. Not surprisingly, almost every investigated anomaly is significantly more present in individuals with NDD - confirming that clinicians should be monitoring the development of children with a congenital anomaly closely.

The subgroup-, EUROCAT- and analyses on enrichment are only some of the numerous examples that are possible with a data set of this size and quality. To further understanding of rare disease, global, open science and data sharing are essential, hence, PhenomAD-NDD is freely available online (Figure 3) on the Human Disease Genes website series (https://humandiseasegenes.nl/phenomadndd/) [37]. The latter also includes a tool allowing clinicians and researchers to calculate the enrichment of clinical features for syndromes of interest.

### 3.1 Conclusion

Pediatric individuals with neurodevelopmental disorders report problems in each organ system, and almost all congenital anomalies are more prevalent in these individuals compared to the general population. The exact prevalence of each HPO term described in our cohort of 51,227 individuals can be found in PhenomAD-NDD, which is available online. Clinicians and researchers can use these data and a developed online tool to determine whether a clinical feature is truly enriched when describing a novel genetic NDD-related syndrome. Finally, we utilize this data to demonstrate that 32% of enriched clinical features in 33 NDD syndromes are currently missing in the clinical synopsis of OMIM.

## Data Availability

The contents of PhenomAD-NDD are freely available online at (https://humandiseasegenes.nl/phenomadndd/).
There, a tool to calculate enrichment of symptoms for your own dataset is available as well.

https://github.com/ldingemans/PhenomAD_NDD_data

## Acknowledgements

We are grateful to the Dutch Organisation for Health Research and Development: ZON-MW grants 912-12-109 (to B.B.A.d.V. and LELMV), Donders Junior researcher grant 2019 (B.B.A.d.V. and LELMV), and Aspasia grant 015.014.066 (to LELMV). The aims of this study contribute to the Solve-RD project (to LELMV), which has received funding from the European Union’s Horizon 2020 research and innovation program under grant agreement No 779257. Multiple authors of this publication are members of the European Reference Network on Rare Congenital Malformations and Rare Intellectual Disability ERN-ITHACA [EU Framework Partnership Agreement ID: 3HP-HPFPA ERN-01-2016/739516] The funder(s) of the study had no role in study design, data collection, data analysis, data interpretation, or writing of the report.

## Author Contributions

The research included worldwide researchers who were involved throughout the research process and each had their own role and responsibility. The study was designed by L.E.L.M.V, B.B.A.d.V and implemented by A.J.M.D, L.E.L.M.V and B.B.A.d.V. The data are owned by the various independent researchers (A.J.M.D, S.J, J.v.R, N.d.L, R.P, J.S.H, B.W.v.B, C.M, C.W.O, M.W, P.J.v.d.S, G.W.E.S, R.F.K, A.T.S.v.S, T.K., D.A.K, L.E.L.M.V, B.B.A.d.V) whereas the intelletual property lies with the leading research group from the Radboud university medical center. The original draft was written by A.J.M.D, L.E.L.M.V and B.B.A.d.V; all authors were involved in reviewing and editing the manuscript.

## 4 Methods

### 4.1 Nijmegen Phenotypic Database

Within the Radboud Biobanks ‘Intellectual Disability’ and ‘Genetics and Rare Disease’ [1], phenotypic and molecular data have been prospectively and systematically captured for individuals with NDD, who were referred to the Radboud university medical center between 2009 and 2021. For all individuals under the age of 18 years, for whom consent allowed, the phenotypes were manually curated and converted into Human Phenotype Ontology (HPO) terms [2]. The medical ethical review board of the Radboud university medical center approved the use of data for the purpose of this study (#2018-4733).

### 4.2 Review of literature

We searched PubMed in February 2021 to find previous studies looking at the associated clinical features of pediatric individuals with NDD. The following search terms were used: mental retardation comorbidity; intellectual disability comorbidity; developmental delay comorbidity; “intellectual disability” AND “congenital anomalies”; “mental retardation” AND “congenital anomalies”; “developmental delay” AND “congenital anomalies”. The titles of the studies resulting from this search were screened for English language, content, and age of included individuals, after which we made a selection. We read the abstracts of these selected studies assessing eligibility, resulting in studies describing the associated somatic comorbidity of a cohort of individuals <18 years with NDD were included (Figure 1, Supplementary table 1).

Since we were interested in the phenotype of the general NDD population, studies focusing on a specific genetic syndrome were excluded. Also studies with a primary focus on psychiatry were excluded — yet, studies where psychiatry was described as comorbidity of NDD were included. We checked the references of all identified articles for possible further inclusions. If this led to additional inclusions, the references of these papers were reviewed as well, ensuring the most complete data set achievable. For every included study, it was checked on which clinical features it reported — as to obtain a realistic denominator for each. For example, if a study reported on ophthalmological symptoms in individuals with NDD, the study was included and used to determine the prevalence of ophthalmological symptoms, but not when calculating the prevalence of for instance congenital heart disease, since these were not assessed.

To provide a homogeneous data set, the clinical data of all papers selected for inclusion were converted to HPO terms. These data were then used to create PhenomAD-NDD: the aggerated data set of the prevalence of all clinical features in individuals with NDD. For assessments of median positively scored HPO terms in all of PhenomAD-NDD parental ancestor terms were included to complete clinical information and to minimize interobserver variability (Figure 2).

In the process of synthesizing data for our meta-analysis, a critical challenge was the inherent limitation of working with summarized data from each study, as we did not have access to the individual-level data. Specifically, we were able to discern the prevalence of individual clinical features (represented as HPO terms) in a given study cohort, but the overlap or co-occurrence of these features within individual participants remained elusive. To address this, while aggregating data, each clinical feature was added as a distinct HPO term. However, given that HPO terms can be nested, with specific symptoms falling under broader categories, there was an inherent risk of inadvertently inflating prevalence figures. To mitigate this, we instituted the safeguard that that the combined prevalence of an HPO term and its related parent or ancestor terms could never exceeded the total patient count from the respective study. Additional mitigations in the synthesis included: i) if a study provided only percentages, the number of patients that had that specific term was calculated using the total number of individuals — and rounded if there was ambiguity; and ii) in cases where a given feature corresponds to multiple HPO terms (for instance, when endocrine and metabolic features were described as one), that feature was excluded.

### 4.3 Subgroup analysis

Subgroup analysis to determine whether sex (male vs. female) and severity of ID (mild/moderate vs. severe/profound) confounded the phenotype were limited to individuals from the Nijmegen Phenotypic Database as this information was not retrievable from the literature included studies. The prevalence of symptoms between subgroups were compared, and statistical significance was assessed using a two-sided chi-squared test with Bonferroni correction for multiple testing, leading to thresholds for significance of *p*<0.05/39 (the number of features investigated)=0.00128, and was *p*<0.05/35 (the number of features investigated in this analysis)=0.00143, for sex difference, and severity of ID, respectively.

### 4.4 Comparison to EUROCAT

We consulted the European network of population-based registries for the epidemiological surveillance of congenital anomalies (EUROCAT) for baseline prevalence of congenital anomaly subgroups in the general population [3–5]. We included all congenital anomalies reported in live births between 2000 and 2016. Only registries that reported data on all congenital anomalies (referred to as “full registries” by EUROCAT) were eligible; Registries in which only a subset was evaluated were excluded for analysis. Two-sided chi-squared tests with Bonferroni correction for multiple testing were used to assess statistical significance, yielding a *p*-value of <0.05/65=0.000769 for significance (65 was chosen as the denominator as that is the number of congenital anomalies investigated).

### 4.5 Enrichment analysis

To demonstrate the power of PhenomAD-NDD, a proof-of-concept study was performed assessing the clinically enriched features for 33 NDD related syndromes. Here, enriched clinical features are defined as symptoms that are significantly more prevalent in a specific genetic disorder than in the general NDD population. The data of these syndromes were collected using the Human Disease Genes website series [6], an initiative to collect phenotypic information in a standardized manner using HPO terms for patients with a molecularly (likely) pathogenic variant (e.g. American College of Medical Genetics and Genomics class 4/5) in the gene of interest, and then compared to the prevalence of that in PhenomAD-NDD. A two-sided Fisher’s exact test was used, in combination with Bonferroni correction for multiple testing (dividing 0.05 by the number of clinical features that are tested for that specific syndrome), to determine statistical significant enrichments. To minimize spurious correlations, the enrichment analysis was performed with different cut-off values for the minimal prevalence (i.e. clinical features with a prevalence lower than the cut-off value in either the syndrome or PhenomAD-NDD were excluded in that specific analysis). A higher threshold leads (by definition) to less included features, that are usually more specific for that genetic syndrome. Furthermore, to ensure a fair comparison, the individuals diagnosed with these syndromes were not included in the data set of PhenomAD-NDD.

The available HPO terms in the clinical synopsis in OMIM [7] were collected for these 33 syndromes as well using the OMIM API, representing the de facto standard of phenotypic information in clinical practice. For all enriched symptoms, it was noted whether the clinical synopsis actually included that specific clinical feature, or whether it was missing. To ensure a fair comparison, all parent-HPO-nodes of all included HPO terms in the clinical synopsis were first made positive.

## Data Availability Statement

The contents of PhenomAD-NDD are freely available online at (https://humandiseasegenes.nl/phenomadndd/). There, a tool to calculate enrichment of symptoms for your own dataset is available as well.

## Code Availability Statement

If the tools provided online are insufficient, or if others would like to implement the enrichment analyses themselves, all code used in this study are available on request from the corresponding author.

## Supplementary data

**Supplementary figure 1:**
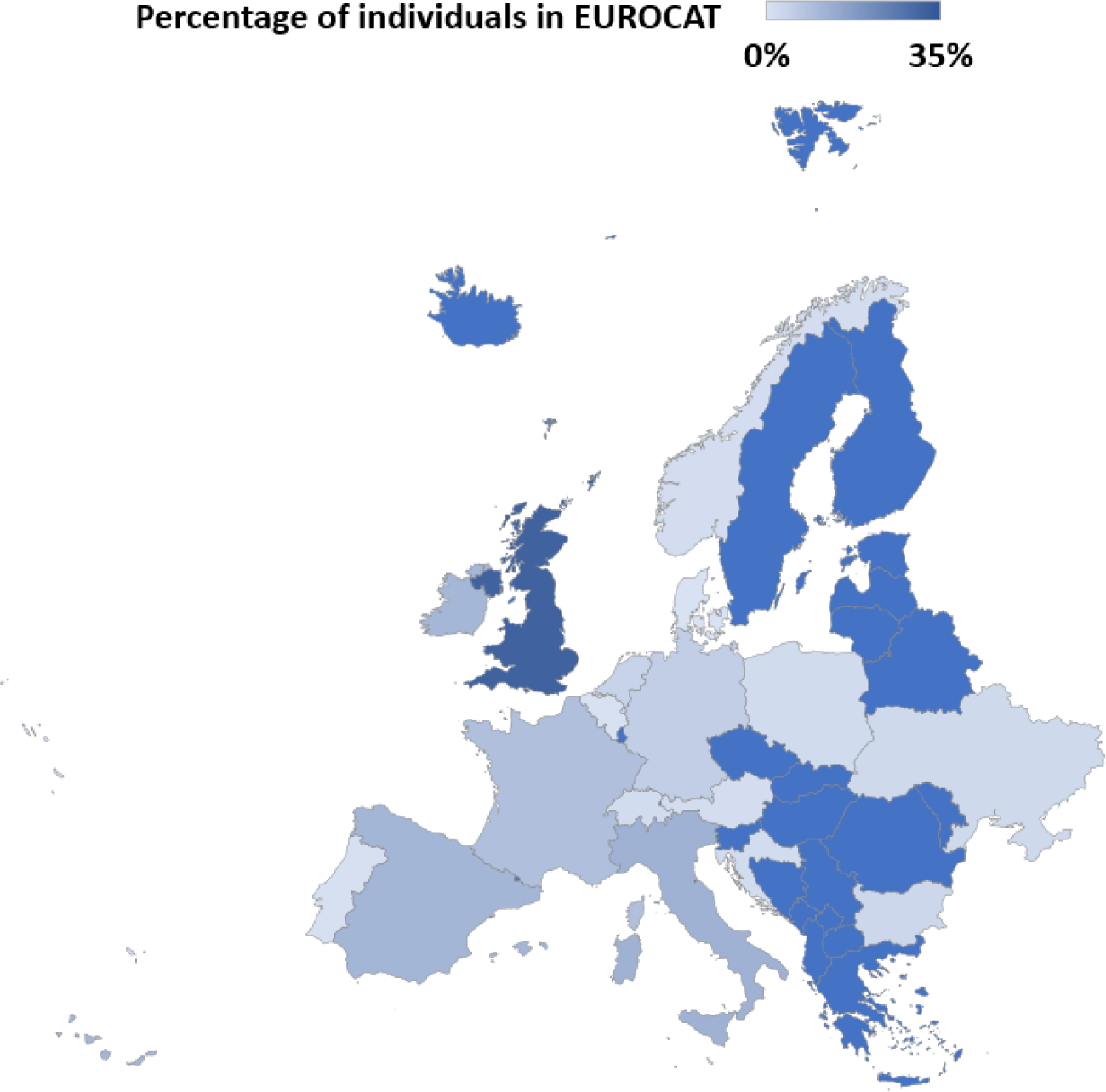
The 18 countries whose institutions contribute data to the EUROCAT registry, with their relative contribution in various shades of blue. Of note, the total number of individuals in the EUROCAT registry is 11,616,332.

**Supplementary figure 2:**
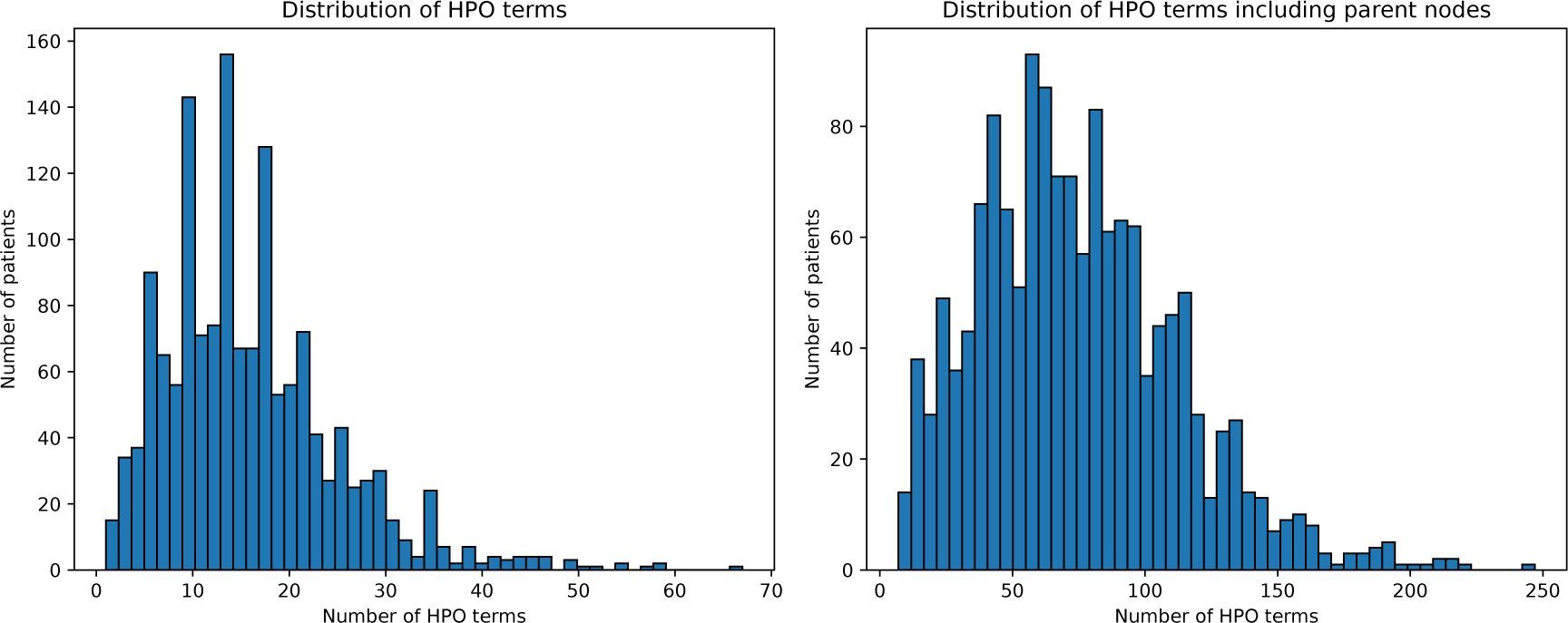
The distribution of the HPO terms of the individuals seen at the outpatient clinic in the Radboud university medical center.

